# Investigation of a Behavioral Interruption Management Strategy on Improving Medication Administration Safety and Efficiency: A Feasibility Study

**DOI:** 10.1101/2023.05.17.23290098

**Authors:** Ginger Schroers, Jill Pfieffer, Dina Tell, Jenny O’Rourke

## Abstract

**Background:** Worldwide, interruptions are pervasive during nurse medication administration and associated with increased frequency and severity of errors. Interruptions also decrease task efficiency which can lead to delayed or omitted patient care. Interruptions cannot always be avoided in healthcare settings; thus, researchers recommend the use of interruption management strategies to mitigate interruptions’ negative effects.

**Aims:** To investigate the feasibility and potential of a behavioral interruption management strategy to mitigate medication errors and improve task efficiency.

**Design:** Multi-methods, two groups, repeated measures, pre-posttest design.

**Methods:** Data were collected January-March 2023. Volunteer undergraduate nursing students were randomly assigned to a control or intervention group. The intervention group received education and training on an interruption management strategy. Quantitative data were collected across three timepoints via direct observation of independent demonstrations of simulated medication administration. The simulated scenarios contained embedded interruptions. Outcomes measured included errors, medication preparation duration, and duration of time to implement the interruption management strategy. Descriptive statistics were analyzed using Microsoft Excel. Qualitative data of participants’ perceptions and use of the strategy were collected via semi-structured interviews.

Thematic analysis was performed.

**Reporting Method:** Equator guidelines were followed using the STROBE reporting method for the observed quantitative data. SRQR guidelines were followed in reporting the qualitative data.

**Results:** Nineteen students participated in the study. Intervention group participants had larger improvements in errors and task durations compared to the control group. Implementation of the strategy averaged four seconds. Participants described the strategy as easy to use and remember, and voiced using the strategy outside of the study.

**Conclusions:** Findings demonstrate that the study-described behavioral interruption management strategy was feasible to teach and implement, and associated with decreased errors and improved task efficiency. Use of the strategy has implications to increase patient safety through improved medication administration safety and efficiency. Future studies are recommended to gain a better understanding of the strategy’s effectiveness.

## Introduction

Interruptions, defined as unplanned breaks during an activity to give attention to a different activity (Brixey et al., 2007), are pervasive in healthcare settings worldwide, particularly to nurses during medication administration (MA) (Schroers, 2018; Zhao et al., 2019). Interruptions are associated with increased frequency (Bonafide et al., 2020; Tsegaye et al., 2020) and severity of medication administration errors (Westbrook et al., 2010) as well as longer nursing task completion times (Cole et al., 2016; Kwon et al., 2021; Schroers et al., 2023). Longer task durations can result in delayed and/or omitted patient care, including failure to administer prescribed medication, thus negatively impacting patient safety.

## Background

Many healthcare settings have implemented efforts, often based on the barrier model, to decrease interruptions during MA. The barrier model involves blocking the source of risk (e.g., interruptions) and what needs protecting (e.g., nurses administering medication) (Gao et al., 2021). For instance, nurses may wear a “do not interrupt” vest or enter a quiet zone when preparing medication. However, these interventions do not consistently decrease interruption and/or error rates (Berdot et al., 2021; Raban & Westbrook, 2014) and some researchers (Schutijser et al., 2019; Westbrook et al., 2017) have raised concerns surrounding their feasibility and sustainability. In addition, Gao et al. (2021) questioned the barrier model’s compatibility with work in hospital environments due to the urgent nature of some interruptions, such as a patient emergency.

Interruptions in healthcare settings are, at times, inevitable. Thus, medical (Thomas et al., 2015) and nurse researchers (Hayes et al., 2015; Schutijser et al., 2019; Westbrook et al., 2017) recommend that healthcare workers and students learn to manage, or handle/respond to, interruptions in ways that can mitigate their negative effects. Behaviors commonly used when a person is interrupted can be categorized as: (a) *Engage - immediately stop the task and give attention to the interruption*, (b) *Multitask - divide attention between the task and interruption,* (c) *Mediate - perform an action to support resumption of the task,* and (d) *Block - ignore the interruption* (Colligan & Bass, 2012).

See Table 1.

**Table 1.** Interruption Management Behavior Operational Definitions (Colligan & Bass, 2012)

| **Interruption Management Behavior-Definition** |
| --- |
| **Engage**- immediately suspend the primary task to give attention to the interruption source |
| **Multitask**- divide attention between the primary task and interruption source; the primary task and the interruption source are similar priorities |
| **Block**- refuse to respond to the interruption source (e.g., ignore or use of hand signals to indicate refusal) |
| **Mediate**- perform an action to support resumption of the primary task before giving attention to the interruption source (e.g., mark the step in the task last completed) |

Nurses primarily engage (Johnson, M. et al., 2017b) and nursing students predominantly engage or multitask (Schroers et al., 2021) when interrupted during MA. Engaging and multitasking during an interruption can lead to errors due to the increase in cognitive demands that occur when switching between tasks. Mediating by forming cues reduces cognitive demands and can decrease errors (Altmann & Trafton, 2002), yet there is scant healthcare research on mediation of interruptions.

Nursing students have expressed a need to learn how to properly manage interruptions (Schroers et al., 2021). Simulation affords a safe environment to practice interruption management during high-risk tasks, such as MA. However, a recent integrative review (Hill et al., 2022) of studies that used simulation to teach participants (including nurses, medical students, and nursing students) interruption management found that most used experiential learning only. In other words, participants were provided the opportunity to feel what it was like to be interrupted but were not taught how to handle interruptions. As Hill et al. (2022) assert, interruption management requires more than the experience of being interrupted - it also requires intentional training and practice. To the authors’ knowledge, only two studies have examined intentional training and practice of an interruption management strategy for use in healthcare settings.

Henneman et al. (2018) pilot tested an interruption management strategy, the Stay S.A.F.E. strategy, with 20 registered nurses using a simulated context. When interrupted during a task, the participants were instructed to **<u>stay</u>** in place, **<u>s</u>**ay aloud the task, **<u>a</u>**cknowledge the interrupting person, **f**ixate on the place in the task, and **<u>e</u>**stimate the time when attention can be given to the person. The strategy was subsequently tested with nursing students during simulated MA scenarios (Vital & Nathanson, 2023). Both nurses (Henneman et al., 2018) and nursing students (Vital & Nathanson, 2023) had a reduction in the time distracted from the primary task after learning the strategy.

## The Study

The purpose of this feasibility study was to inform a future larger study by investigating a researcher developed interruption management education bundle that included a revised version of the Stay S.A.F.E. strategy (Henneman et al., 2018). The study aims were to explore participants’ (a) implementation and (b) perceptions of the strategy, and to describe changes across three timepoints of participants’ (c) interruption management behavior, (d) medication administration error frequencies, and (e) duration of medication preparation during interrupted simulated MA.

## Theoretical Frameworks

### Memory for Goals Model

The Memory for Goals (MFG) cognitive science model (Altmann & Trafton, 2022) underpinned the interruption management bundle. The model asserts that associative cues (i.e., retrieval cues) must be formed during interrupted tasks to facilitate task resumption. Cues can be environmental and/or in the person’s mental context. When a person notices an alert (e.g., phone ringing) a cue should be formed during the interruption lag before stopping the task and giving attention to the interruption source. To put it another way, mediation should be used. When the interruption ends, the cue should be retrieved to aid in task resumption. According to the model, associative cueing can improve task efficiency and accuracy by bringing the person back to the proper step in the primary task without omitting or repeating steps (Altmann & Trafton, 2002). Evidence outside of healthcare provides support for the use of cues during interrupted tasks (Dodhia & Dismukes, 2009; Gartenberg et al., 2014; McDonald & Durso, 2015). See Figure 1.

**Figure 1.**
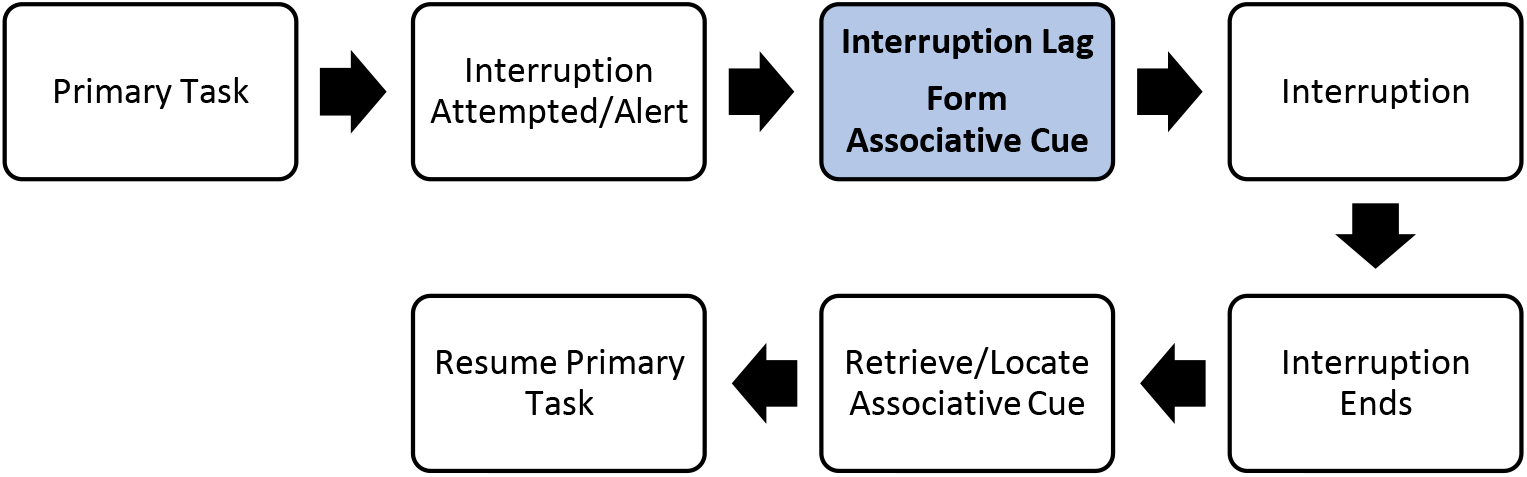
Use of Associative Cueing during an Interrupted Task *Note.* This illustration depicts the steps of when to form an associative cue during an interrupted task.

### Event Segmentation Theory

Event Segmentation Theory (EST) (Zacks et al., 2007) guided the timing of the embedded interruptions during the simulated pre/posttest scenarios. According to EST, tasks can be divided into coarse and fine segments. A coarse segment of medication preparation, for example, could be completing preparation of one of multiple scheduled medications. Fine segments could be any subtask of medication preparation, such as verifying the label of a medication or drawing up medication into a syringe. Prior research demonstrated that interruptions during subtasks/fine segments increased self-reported time pressure and mental demand among study participants compared to when interrupted during coarse segments (Adamczyk & Bailey, 2004). Interruptions during fine segments are believed to increase the risk for errors and time to resume the primary task (Powers & Scerbo, 2023), thus were chosen for the timing of the interruption in this study. The timing of the embedded interruptions during a fine segment of medication preparation provided a more accurate assessment of the association between the study independent variable - the interruption management strategy - and dependent variables of errors and task durations.

## Methods

### Research Design

A multi-methods, two group, repeated measures, pre-posttest study design was used. Data were collected January-March 2023. STROBE guidelines were followed in the reporting of observations (Von Elm et al., 2007) and Standards for Reporting Qualitative Research (SQRQ) guidelines were followed in reporting the qualitative findings (O’Brien et al., 2014). Participants were randomly assigned to a control or intervention group. Both groups received a MA education bundle; the intervention group also received an interruption management bundle. Control group participants were offered the interruption management education after data collection was completed. Participants were blinded to the specific study aims to minimize changes in participant behaviors that could have distorted the findings (Polit & Beck, 2021).

### Participants and Setting

A convenience sample of undergraduate accelerated Bachelor of Science in Nursing (ABSN) students was recruited from a Midwest United States private university. Email invitations were sent to a total of 77 eligible students, and recruitment flyers were posted in common areas on campus. The recruitment period began in November 2022 and ended in January 2023. Inclusion criteria were enrollment in the second semester of the ABSN program, and a passing grade in an introductory nursing skills course taken in the first semester. Students learn, practice, and must demonstrate MA competency (in a lab setting) to receive a passing grade in the introductory skills course. Thus, this criterion ensured participants had baseline MA knowledge and skills, and applied control for participants’ MA experiences as a potential confounding factor. Recommendations for feasibility sample sizes vary from 10-12 to 60-75 per group (Lewis et al., 2021). The research team aimed to enroll 10-12 participants per group (20-24 total). The study setting was the university’s simulation center.

### Procedures

Students participated in the study on two days during one semester. Study day 1 consisted of a group of 4-6 participants over a 3.5 hour period. On study day 1, participants 1) completed an individual pretest, 2) received an education bundle as a group, and 3) completed an individual posttest. The pre and posttests were independent demonstrations of interrupted simulated MA. Study day 2 was an individual one hour appointment that consisted of a second posttest and semi-structured interview. Study day 2 occurred 4-8 weeks after study day 1.

### Education Bundles

The principal investigator (PI) developed two educational bundles for the study. The bundles included didactic education, expert role modeling via video demonstration, and MA practice in a simulated context. Three nurse educators/content experts employed by three different organizations assessed the content validity (CV) of the bundles for relevance to improving learners’ ability to safely administer medication. The I-CVI and S-CVI of both bundles scored 1.00, thus were judged as having excellent CV (Polit & Beck, 2021). The MA bundle focused on medication safety principles (e.g., verifying the medication dose) that participants’ learned in the preceding semester. All participants received identical MA bundles.

#### Interruption Management Bundle

The interruption management bundle was guided by the MFG model (Altmann & Trafton, 2002), findings of nursing students’ use of cues during interrupted simulated MA (Schroers et al., 2021), experts’ recommended strategies for novice nurses (Johnson, K. et al., 2021), and Henneman et al.’s (2018) Stay S.A.F.E. strategy. The interruption management bundle included a discussion and video demonstration the Stay S.A.F.E. strategy during interrupted MA. Examples of cues for use when mediating an interruption were provided that included holding medications, mentally stating the last step in the task completed, and placing a finger on the working area.

The PI adapted Henneman et al.’s (2018) strategy to be used with various sources of interruptions, not only a person, such as a non-urgent alarm or phone ringing. Placement of mediation with cues was also moved earlier in the revised strategy, during the interruption lag, to align with the MFG model (Altmann & Trafton, 2002). In addition, the “E” in Stay S.A.F.E. was changed from “estimate time…” to “evaluate the urgency of the interruption”.

Researchers (Johnson, K. et al., 2021) assert that nurses need to quickly and efficiently determine the urgency of an interruption to decide what takes priority - the interruption or the current task. Examples of ways to evaluate the urgency of an interruption caused by a person were provided, and included direct questions, such as “Is this urgent?” and assessment of the interrupting person’s body language. See Table 2.

**Table 2.**
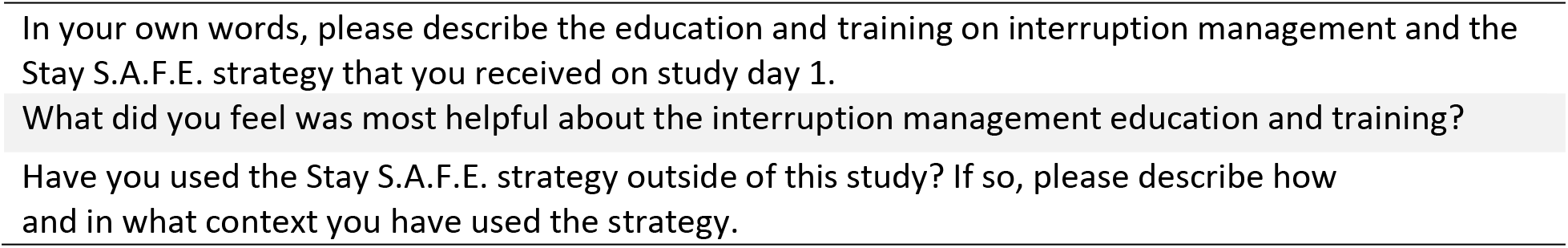
Revised Stay S.A.F.E. Strategy (Adapted from Henneman et al., 2018)

### Simulated Scenarios

The design of the pre and posttest scenarios followed the Healthcare Simulation Standards of Best Practice^TM^ (Watts et al., 2021). Scenarios were comparable in complexity and number (*n* = 2) of medications ordered during the simulated time. The case components (*n* = 13) (e.g., scripts; provider orders) and entire cases were assessed by six simulation experts and judged to have excellent CV. The I-CVI and S-CVI were calculated as 1.00 (Polit & Beck, 2021). All participants experienced identical simulated scenarios. Further details of the simulated scenarios and educational bundles are available upon request by contacting the first author.

#### Interruptions

A trained actor was instructed to cause one interruption to the participant in the medication room when the participant was reading a medication label - a fine segment/subtask of medication preparation (Zacks et al., 2007). Before the pretest/posttests, participants were introduced to the actor and informed that they were playing a team member role during the simulated scenarios, however, participants were not told that the actor would cause an interruption. The same actor played a nurse role in the pretest and first posttest on study day 1; the interruption was a conversation about workflow. A different actor played a physician role in the second posttest on study day 2; the interruption was a request for patient information participants were known to have received during prebriefing (e.g., most recent vital signs). The sources, reasons, and location for the interruptions were guided by observational evidence of interruptions to nurses during MA (Johnson, M. et al., 2017a).

### Data Collection and Analysis

Quantitative data were collected by two independent researchers via direct observation. The researchers collected data in real-time, from a control room that provided live visual and audio of patient rooms, and audio-video recordings. Audio-video recordings were required to gather data from the medication room (this room was unable to be viewed in real-time), confirm participant actions in patient rooms, and note time durations.

Data were documented on a pre-existing validated tool (Johnson, M. et al., 2017a) that included checkboxes to mark if a medication administration error occurred (YES/NO), type of error, and interruption management behavior used by the participant. Interrater reliability (IRR) of the medication administration error definitions have reported kappa coefficients of 0.94-0.96 (Westbrook et al., 2010). The interruption management behavior definitions have reported CV scores of 0.80-1.00 (Schroers et al., 2021). Some behaviors were expected to be difficult or unable to observe, such as mediation with a cue, therefore two methods were used to collect these data. After the pre and posttests, written and verbal definitions of interruption management behaviors (see Table 1) were provided to participants, and participants noted the behavior they used on a researcher made form. The self-reported behavior was compared with the noted observation to verify or correct these data.

The PI added an area to the instrument to document durations of time (in seconds) participants (a) spent in medication preparation, (b) stopped medication preparation to give attention to the interruption source, and, when applicable, (c) mediated the interruption using the revised Stay S.A.F.E strategy. The audio-video footage provided timestamps that enabled accurate notation of start and end times. Similar to methods used by Cole et al. (2016), Kwon et al. (2021), and Schroers et al. (2023), the duration of time a participant stopped and/or mediated the interruption was subsequently subtracted from the medication preparation time to provide a more accurate reflection of the interruptions’ effect on task duration.

Prior to study initiation, the data collectors familiarized themselves with the operational definitions (Tables 1 and 3), data collection instrument, and indications for noting the time durations. The researchers independently collected data on five random cases (25% of all cases) to establish IRR. Using percent agreement (McHugh, 2012), IRR across 184 items/data points was calculated as 99%. Descriptive statistics were analyzed using Microsoft 365 Excel software.

**Table 3.** Operational Definitions of Medication Administration Errors

|  |
| --- |
| **STAY** physically where you are. |
| Stay engaged with the task. Continue to hold any items you are working with when possible. |
| Associate cues. |
| Fixate on your place in the task for 1-2s and mentally say to yourself what you are doing.<br>Place your **F**inger on the area you are working. **F**ind a natural break in the task when you can pause. |
| Evaluate the urgency of the interruption. If urgent, tend to the interruption source immediately. If non-urgent decide if you can ignore or delay. If decide to delay, finish any critical safety sub-tasks before giving attention to the interruption. |

After the second posttest, intervention group participants rated the ease of use and remembering the revised Stay S.A.F.E. strategy by viewing a 4-point Likert scale (*1 = not easy, 2 = somewhat easy, 3 = very easy, 4 = extremely easy*), and participated in an individual 10-20 minutes recorded semi-structured interview. Using a descriptive qualitative approach (Polit & Beck, 2021), the PI conducted the interviews to explore participants’ perceptions and use of the interruption management education/strategy. See Table 4.

**Table 4.**
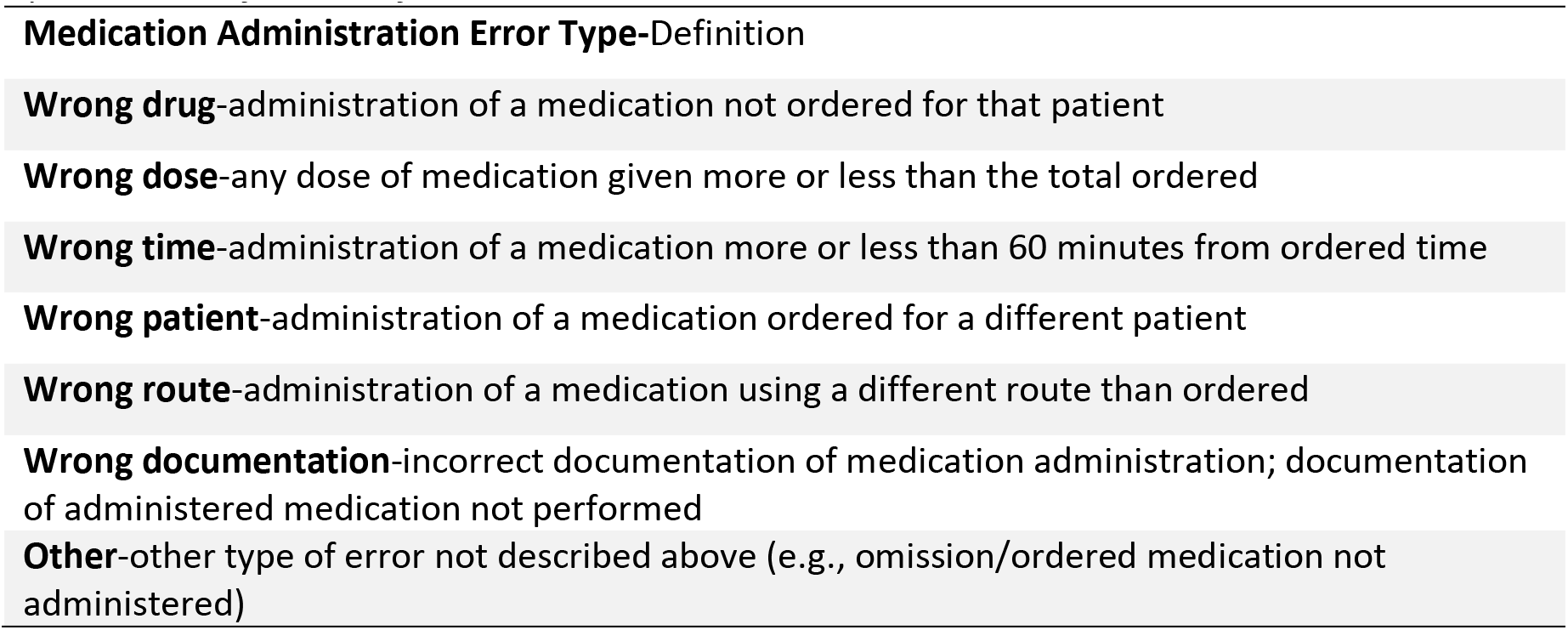
Examples of Semi-Structured Interview Questions

Recordings of the interviews were transcribed by a professional transcription service and checked for accuracy by the PI. Two research team members independently performed thematic analysis (Vaismoradi & Snelgrove, 2019) of the qualitative data by iteratively reading the transcripts, creating broad codes, and potential themes. The team members discussed the identification and fittingness of data to the themes until a consensus was reached. To enhance trustworthiness of the qualitative findings, three participant member checks were conducted to ensure credibility and exemplars are provided to enhance confirmability (Polit & Beck, 2021).

### Ethical Considerations

The study site provided Institutional Review Board (IRB) approval. The study was voluntary, conducted outside of class time, and did not impact any student course grades. No research team members were current instructors of any participants. Participants signed informed consent which included consent to be audio-video recorded during the simulated scenarios and interview sessions. Only the research team members were able to view participants when they completed the pre/posttests. Participants received a $50 Amazon gift card as appreciation for their time in the study.

## Results

### Participants/Sample

Twenty-one students enrolled in the study. Two students withdrew their enrollment on the morning of study day 1 due to personal reasons. A total of nineteen students participated in study day 1; 18 participated in study day 2. One intervention group participant did not return for study day 2 for unknown reasons. Participants’ age in years ranged from 22-54 with a mean age of 28.7 and median age of 26. Most students (79%) had past or current healthcare work experience. Participants’ self-reported grade point averages (GPA) were comparable among groups: control group mean GPA was 3.49; intervention group mean GPA was 3.45. No participants received specific interruption management training prior to the study.

### Interruption Management

During the pretest, control and intervention group participants engaged or multitasked during the interruption. During the two posttests, nearly all (94%) intervention group participants mediated the interruption. Three control group participants mediated the interruption in posttest 2 by placing and keeping their finger on the working area. See Table 5.

**Table 5.**
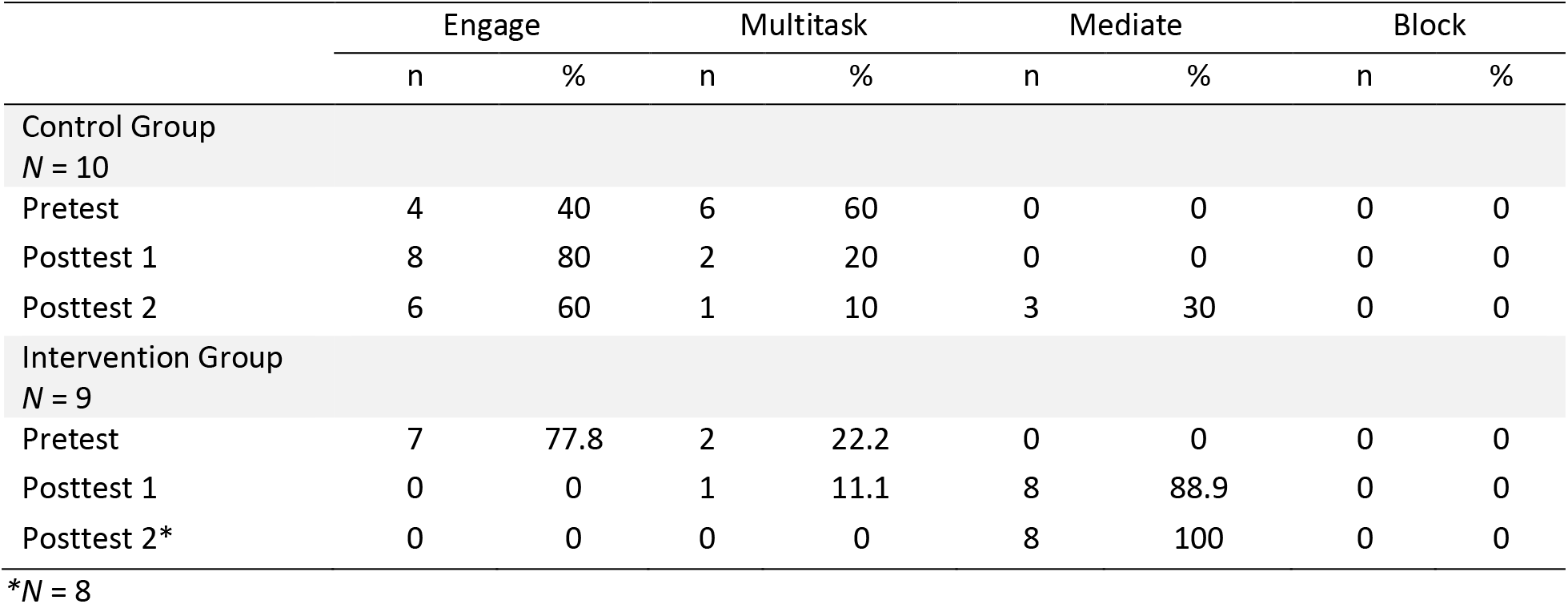
Interruption Management Behavior used by Participants

### Stay S.A.F.E. Strategy Implementation

Eight out of nine (88.89%) intervention group participants implemented the revised Stay S.A.F.E. strategy during posttest 1; 100% (*n* = 8) used the strategy in posttest 2. Implementing the strategy had a mean duration of 5s (SD:5.06s) in posttest 1, 2.5s (SD:3.07s) in posttest 2, and 4s (SD:4.23s) across both posttests. See Figure 2.

**Figure 2.**
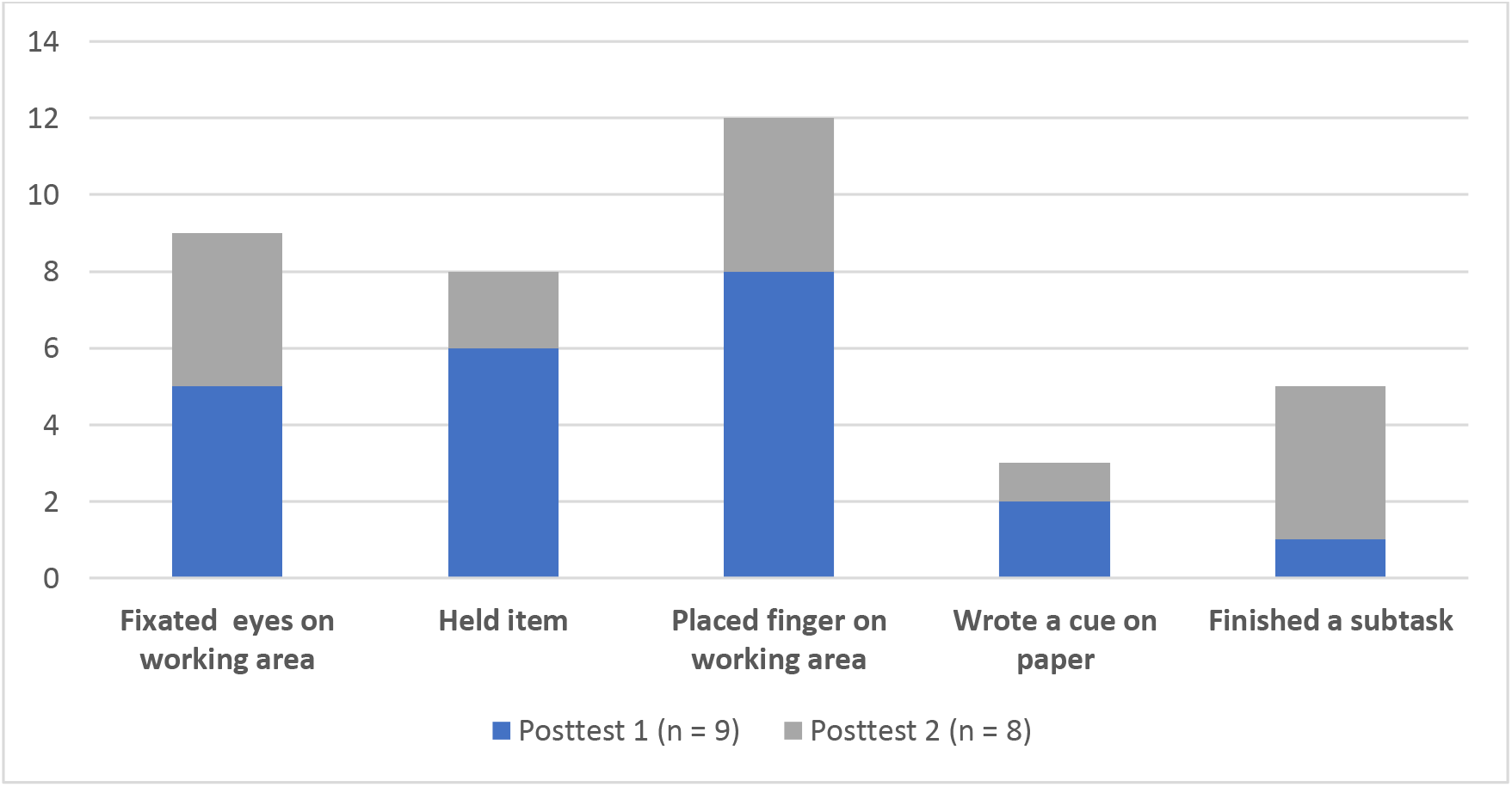
Frequencies of Stay S.A.F.E. actions used *Note.* This chart illustrates the frequencies of specific actions/cues used among intervention group participants during the posttests.

### Medication Administration Errors and Durations

Medication administration errors decreased among both groups from the pretests to posttests. The intervention group had larger percentage decreases compared to the control group. Task durations decreased from pretest to posttest 1 among both groups, but slightly increased among both groups from pretest to posttest 2. See Tables 6 and 7.

**Table 6.** Medication Administration Error Frequency Percent Change

| **Percent Change** | **Pre -**<br>**Posttest 1** | **Pre-**<br>**Posttest 2** |
| --- | --- | --- |
| Control Group | 62.5%<br>decrease | 75%<br>decrease |
| Intervention Group | 75%<br>decrease | *86.2%<br>decrease |

*Note.* \*Excludes pretest data from one intervention group participant who did not complete posttest 2.

**Table 7.** Medication Preparation Duration Percent Change

| **Percent Change** | **Pre-**<br>**Posttest 1** | **Pre-**<br>**Posttest 2** |
| --- | --- | --- |
| Control Group | 14.9%<br>decrease | 2.9%<br>increase |
| Intervention Group | 16.4%<br>decrease | *1.8%<br>increase |

*Note.* \*Excludes pretest data from one intervention group participant who did not complete posttest 2.

### Participant Perceptions and Use of the Strategy

Three themes emerged through the qualitative analysis of the interviews with the intervention group participants: the strategy (a) is easy to use and remember, (b) increased knowledge and confidence in handling interruptions, and (c) has utility in different contexts.

### Easy to Use and Remember

All participants in the intervention group (*n* = 8; 100%) rated the revised Stay S.A.F.E. strategy as “extremely” or “very” easy to use. Most (75%) rated the strategy “extremely” or “very” easy to remember.

### Knowledge and Confidence

When asked what was most helpful regarding strategy, one participant stated “Well, first, learning about it….we didn’t even know that [interruption management] was a thing until you told us.” Others voiced it was helpful to be told “specifically” what they could do when interrupted and that the education and training gave them “confidence” in handling interruptions.

### Utility in Different Contexts

Most (75%) intervention group participants reported using the strategy outside of the study. One participant described being in their clinical rotation, and while programming a patient’s IV pump, a family member began asking the student questions: “…I first just put the pump to what I think it should be set to and put my finger on it, so when I get back, okay, I was focusing on the rate being correct. And then I just glanced at them… and I was like, ‘Just give me one moment please,’… I didn’t move.” Others discussed using the strategy when studying: “If I’m studying with somebody, and they’re interrupting me while I’m reading… if it’s on my iPad, I’ll mark it on there where I left off…”

## Discussion

Prior to receiving the interruption management education and training, intervention group participants multitasked or engaged when interrupted. Multitasking and engaging can increase risks for errors, thus are not recommended for use during high-risk tasks. It is recommended that healthcare students and workers learn to use strategies that can mitigate risks when interrupted, yet there is limited evidence available to inform strategies.

To the authors’ knowledge, only two prior studies of teaching an interruption management strategy for use in healthcare have been conducted. Henneman et al. (2018) investigated the original Stay S.A.F.E. strategy with nurses, however examined different outcomes than this study. Vital and Nathanson (2023) investigated the original Stay S.A.F.E. strategy with nursing students and, like this study, examined errors and task durations.

### Medication Administration Errors

This study found a larger percent decrease in the number of medication administration errors among the intervention group compared to the control group. Vital and Nathanson (2023) reported no significant differences in error frequencies between their intervention group (who received education of the original Stay S.A.F.E. strategy) and control group. It is possible that the revisions made to the Stay S.A.F.E. strategy in this study contributed to the larger improvements in errors among intervention group participants, however, these findings should be taken with caution as this was the first study conducted with the revised strategy.

### Medication Preparation Duration

The intervention group in this study had a larger decrease in task duration from the pretest to posttest 1, consistent with Vital and Nathanson (2023) who reported shorter MA completion times among their intervention group. Both groups in this study, however, had small increases in task durations from the pretest to posttest 2. The increases may have been due to the first posttest occurring on study day 1, which was the same study day participants pretested and received education and practice of interrupted MA. Students likely had better recall of the education and practice during the first posttest test, which may have influenced the completion times (i.e., carryover/practice effect).

However, it is important to re-iterate that the increased time to complete medication preparation was small. There was less than a 2% increase among the intervention group, suggesting that participants retained much of what was learned and practiced on the first study day. Knowledge and skill retention was also supported by the decrease in errors across time points and use of the Stay S.A.F.E. strategy by intervention group participants in the second posttest.

### Implementation of the Strategy

The revised Stay S.A.F.E. strategy was feasible for nursing student participants to use, and participants cited many benefits of the strategy. Participants implemented the strategy using various and multiple actions/cues, yet the averaged time using the strategy was only four seconds. In addition, participants reported the strategy as easy to use and remember, and stated that it was helpful to be given specific instructions on how to handle interruptions.

Students articulated that the education and practice of the strategy gave them the confidence to delay (by mediating) their attention to a non-urgent interruption to focus on MA safety. This was an important finding as nursing students report that they lack confidence in handling interruptions and are unsure of how to maintain safety when interrupted during MA (Schroers et al., 2021). Interestingly, three control group participants, who did not receive the interruption management education, nor did they mediate the interruption in the pretest or first posttest, mediated the interruption in the second posttest. It is unknown what influenced their use of mediation.

### Challenges

The primary challenge the researchers faced during the pre/posttests was timing the interruption to occur when participants were reading a medication label. The actor was physically located in the hallway, about 3 feet from the entrance to the medication room - the medication room door was ajar when participants entered the room. However, during two posttests a participant unexpectedly closed the medication room door, causing the actor to need to estimate when to create the interruption. To address this challenge in future studies, it is recommended to position the actor inside the medication room with a clear visualization of the participants’ actions.

### Strengths/Limitations

Strengths of this study were the rigorous methods used in the research design, including the use of psychology theories to guide the education bundle, revised strategy, and placement of the interruption during the simulated scenarios. Excellent CV was obtained on the education bundles and simulated scenarios, and high IRR was obtained between the two data collectors. Limitations include this being a single-site feasibility study thus limiting the generalizability of the findings. In addition, a potential for selection bias was present. Participants volunteered for the study therefore may have been more confident in their MA and/or simulation skills, resulting in better MA performance compared to the general nursing student population.

### Recommendations and Implications for Practice

Future research with different populations and contexts, and use of various interruption sources (e.g., phone ringing, family member) is recommended to gain a better understanding of the strategy’s effectiveness. It is also recommended that educators and other healthcare leaders educate employees and students on the dangers of interruptions, and strongly discourage non-urgent interruptions to colleagues during high-risk tasks. Decreasing non-urgent interruptions, and implementing evidence-based strategies to handle interruptions when they do occur, can lead to improvements in patient safety and healthcare quality.

## Conclusion

This study’s findings contribute to a gap in the healthcare literature of evidence for interruption management. The revised Stay S.A.F.E. strategy was found to be feasible, associated with decreased errors and improved task efficiency, can be applied in a variety of contexts among different disciplines. Interruptions will never completely cease in healthcare settings; thus, it is critical that healthcare students and workers learn to appropriately manage interruptions in ways that can mitigate their negative effects. The authors are planning a future, powered, multi-site study using the same methods with changes to the physical location of the actor and addition of various interruption sources that will include people and devices.

## Data Availability

All data produced in the present study are available upon reasonable request to the first author.

## Acknowledgements

Acknowledgement of funding support

Loyola University Chicago Marcella Niehoff School of Nursing provided financial support for this study.

